# Association of close-range contact patterns with SARS-CoV-2: a household transmission study

**DOI:** 10.1101/2022.12.22.22283843

**Authors:** Jackie Kleynhans, Lorenzo Dall’Amico, Laetitia Gauvin, Michele Tizzoni, Lucia Maloma, Sibongile Walaza, Neil A. Martinson, Anne von Gottberg, Nicole Wolter, Mvuyo Makhasi, Cheryl Cohen, Ciro Cattuto, Stefano Tempia, the SA-S-HTS Group

## Abstract

**Background:** Households are an important location for severe acute respiratory syndrome coronavirus 2 (SARS-CoV-2) transmission, especially during periods where travel and work was restricted to essential services. We aimed to assess the association of close-range contact patterns with SARS-CoV-2 transmission.

**Methods:** We deployed proximity sensors for two weeks to measure face-to-face interactions between household members after SARS-CoV-2 was identified in the household, in South Africa, 2020 - 2021. We calculated duration, frequency and average duration of close range proximity events with SARS-CoV-2 index cases. We assessed the association of contact parameters with SARS-CoV-2 transmission using mixed effects logistic regression accounting for index and household member characteristics.

**Results:** We included 340 individuals (88 SARS-CoV-2 index cases and 252 household members). On multivariable analysis, factors associated with SARS-CoV-2 acquisition were index cases with minimum C_t_ value <30 (aOR 10.2 95%CI 1.4-77.4) vs >35, contacts aged 13-17 years (aOR 7.7 95%CI 1.0-58.2) vs <5 years and female contacts (aOR 2.3 95%CI 1.1-4.8). No contact parameters were associated with acquisition (aOR 1.0 95%CI 1.0-1.0) for all three of duration, frequency and average duration.

**Conclusion:** We did not find an association between close-range proximity events and SARS-CoV-2 household transmission. It may be that droplet-mediated transmission during close-proximity contacts play a smaller role than airborne transmission of SARS-CoV-2 in the household, due to high contact rates in households or study limitations.

**Funding:** Wellcome Trust (Grant number 221003/Z/20/Z) in collaboration with the Foreign, Commonwealth and Development Office, United Kingdom.

## Introduction

South Africa has experienced five waves of severe acute respiratory syndrome coronavirus 2 (SARS-CoV-2) infection, with over 4 million laboratory-confirmed cases by August 2022 ^1^. The true burden is highly underestimated, since based on seroprevalence data, after the third wave of infection, 43% to 83% of the 59.5 million South African inhabitants had already been infected, varying by age and setting ^2,3^.

SARS-CoV-2 transmission is mainly via the respiratory route, with droplet-mediated transmission thought to be the most important but airborne transmission also occurs ^4,5^. Infection from contaminated surfaces has also been described ^4^. Although infection risk is highest from symptomatic individuals ^6^, with the most infectious period one day before symptom onset ^4^, asymptomatic individuals can still transmit SARS-CoV-2 ^7,8^. Households are a focal point for SARS-CoV-2 transmission ^9,10^, especially during peaks of non-pharmaceutical intervention (NPI) restrictions, when movement outside of the household was limited ^9^. Transmission within households can in turn lead to spill over to the community ^11^.

Prior to the widespread availability of SARS-CoV-2 vaccines, most countries relied on NPIs to reduce the transmission of the virus, including wearing face masks, social and physical distancing. While mobility and contact survey data showed that the implementation of NPIs led to a reduction in community contacts ^9,12^ and in turn opportunity for infection, it is still unknown what the role of contact patterns are in the transmission of SARS-CoV-2 in the household. Most analysis relating contact patterns and SARS-CoV-2 transmission done to date has been based on low resolution data collected from contact tracing ^13^, mobility data ^9^ and contact surveys ^12^. To obtain high-resolution contact data, devices broadcasting and receiving radio frequency waves can be used to measure the frequency and duration of close-proximity contacts. This has been used previously to collect contact data in among others, schools ^14^, workplaces ^15^, hospitals ^16^ and households ^17^, which can, in turn, be used for modelling disease transmission. Specifically, for SARS-CoV-2 so far, high-resolution contact data were collected on cruise ships to identify areas of high contact, and to investigate the usefulness of NPIs ^18^.

Understanding the drivers of SARS-CoV-2 transmission in the household, especially contact patterns, can help inform NPIs for future SARS-CoV-2 resurgences and potentially future emerging pathogens with pandemic potential. We aimed to assess the association of household close-range contact patterns with the transmission of SARS-CoV-2 in the household using proximity sensors deployed after the identification of SARS-CoV-2 in the household.

## Methods

### Screening, enrolment and follow-up

We nested a contact study within a case-ascertained, prospective, household transmission study for SARS-CoV-2, implemented in two urban communities in South Africa, Klerksdorp (North West Province) and Soweto (Gauteng Province) from October 2020 through September 2021. Sample size calculations were performed for the main study, but not the nested contact study. For the main study we aimed to assess a significant difference in the household cumulative infection risk (HCIR) between household contacts exposed to SARS-CoV-2 by a HIV-infected vs HIV-uninfected index case for a 95% confidence interval and 80% power. The resulting total sample size was 440 exposed household members. Detailed sample size calculations and methods for the main study have been reported previously ^19^. In short, symptomatic adults (aged ≥18 years, symptom onset ≤5 days prior) consulting at clinics were screened for SARS-CoV-2 with real-time reverse transcription polymerase chain reaction (rRT-PCR) on nasopharyngeal swabs. We enrolled household contacts of SARS-CoV-2 infected individuals identified through screening (presumptive index) with ≥2 household contacts of whom none reported symptoms prior to index case onset. We visited enrolled households three times a week to collect nasal swabs and data on symptoms and healthcare seeking. At enrolment household characteristics (household size, number of rooms used for sleeping, smoking inside the household and household income) and individual characteristics (demographics, education, employment, smoking, HIV infection, underlying illness, if SARS-CoV-2 index case was main caregiver, or sleeping in same room as index case) were collected. Nasopharyngeal (screening) and nasal swabs (follow-up) were tested for SARS-CoV-2 on rRT-PCR using the Allplex™ 2019-nCoV kit (Seegene Inc., Seoul, South Korea) and the first positive of each infection episode was characterised using the Allplex™ SARS-CoV-2 Variants I and II PCR assays (Seegene Inc., Seoul, Korea) and through whole genome sequencing on the Ion Torrent Genexus platform (Thermo Fisher Scientific, USA). We classified the infection episodes as Alpha, Beta, Delta, non-Alpha/Beta/Delta or unknown variant where we were unable to classify the sample as a variant of concern due to primary testing done elsewhere, low viral load or poor sequence quality. Households with multiple SARS-CoV-2 variants circulating at the same time (mixed clusters) were excluded from the analysis. We also collected serum at the first and final household visit for serological testing, using an in-house ELISA to detect antibodies against SARS-CoV-2 spike protein ^20^ and nucleocapsid protein using Roche Elecsys anti-SARS-CoV-2 assay. Individuals were considered seropositive if they tested positive on either assay. Individuals sero-positive at the start of follow-up with no rRT-PCR confirmed SARS-CoV-2 infection during follow-up were excluded from the analysis as they may have been protected from infection _21_.

### Contact pattern measurements

At the first or second visit during follow-up, we deployed wearable radio frequency (RF) proximity sensors ^15^ for two weeks to measure close-range interactions (<1.5 meters) between household members. The proximity sensors exchange low-power radio packets in the ISM (Industrial, Scientific and Medical) radio band. Exchange of packets and Received Signal Strength Indicator (RSSI), suitably thresholded, are used to assess proximity between the devices. A contact interval between two devices is defined as a sequence of consecutive 20-second intervals within which at least one radio packet was exchanged. Each sensor had a unique hardware identifier that was linked to participant study identifiers. Sensors were worn in a PVC pouch either pinned to clothing on the chest, or on a lanyard around the neck based on participant preference. We requested participants to wear the device while at home, to store them separately from other household member sensors at night, and to complete a log sheet every day for the periods the sensors were put on and taken off. During each household visit during the sensor deployment period, field workers confirmed sensors were worn. A deployment log was completed for each household to link the sensor identifier to the participant identifier and to log the date and time sensors were deployed and collected. After sensor collection, batteries were removed to prevent further package exchange between sensors. Sensors were transported to the study office where each sensor was connected to a computer and data downloaded.

### Data analysis

We assumed the first individual with COVID-19 compatible symptoms in the household (individual screened at clinic) was the index case. Any household member testing positive for SARS-CoV-2 within the two weeks from the last positive result for the index case was considered a secondary SARS-CoV-2 case. Contact event data were cleaned using an automated pipeline. We excluded any close-range proximity events outside of the deployment period and that occurred during a 5-minute time slice that the accelerometer did not detect any movement of the sensor. Due to a technical error, some sensors at the Klerksdorp site did not have a valid time stamp and needed additional processing to align the time series of close-range proximity events. This was achieved by computing, for each pair of tags X and Y, the temporal shift that maximizes the correlation between the time series of the number of packets per unit time transmitted by X and received by Y, and the reciprocal time series of the number of packets per unit time transmitted by Y and received by X (operation that can be efficiently carried out working in the frequency domain via Fourier transformation). This allowed us to build a temporal alignment graph between sensors and – as long as there was at least one sensor with a valid timestamp in the household – to use such graph to propagate the valid timestamp to all other sensors, thus recovering global temporal alignment. For the analysis, we only considered close-range proximity events that occurred one day after deployment and one day before collection. Where no timestamp was available, we used data collected from one to ten days after deployment.

We assessed three contact parameters: 1) duration (median daily cumulative time in contact in seconds), 2) frequency (median daily number of contacts with the index/infected individuals over the deployment period) and 3) average duration (cumulative time in contact divided by the cumulative number of close-range proximity events over full deployment period). Median values were preferred over mean values due to the rightly skewed data, and the different number of days with measured contact data for each household after data cleaning. We assessed contact parameters in two ways: 1) median number of close-range proximity events with the presumptive index case and 2) median number of close-range proximity events with all SARS-CoV-2 infected household members (as confirmed by rRT-PCR). The latter assessment was to take into account that the transmission could have been from any of the infected household members, and not necessarily the index case, or that the index case was misclassified.

We constructed contact matrices by combining the median duration and frequency of close-range proximity events for all participants between each age group, respectively. To normalize the matrix based on number of participants, we divided the cumulative contact duration and frequency by the total number of individuals in the two age groups being investigated in each cell.

We assessed the association of contact parameters with SARS-CoV-2 household transmission using the Wilcoxon rank-sum test (considering p<0.05 as significant) and through logistic regression controlling for individual characteristics associated with transmission. To assess factors associated with SARS-CoV-2 household transmission, we performed logistic regression with a mixed effects hierarchical regression model to account for household- and site-level clustering. For the analysis with a defined index case (i.e. investigating close-range proximity events with all presumptive index cases, first person with COVID-19 symptoms), we included only household contacts with their SARS-CoV-2 infection status as the outcome, assessing both index (transmission) and contact (acquisition) characteristics. For the analysis with no defined index case (i.e. investigating close-range proximity events with all SARS-CoV-2 infected household members), we included all enrolled household members (originally considered presumptive index and household contacts), assessing only their own characteristics. For the analysis with close-range proximity events with all SARS-CoV-2 infected household members, we included an offset term in the model to account for the number of SARS-CoV-2 infected members in contact with (number of nodes). We first built the model using individual characteristics to assess factors associated with SARS-CoV-2 transmission (excluding contact parameters). We included age and SARS-CoV-2 variant *a priori*, and assessed other co-variates on univariate analysis, keeping those with p<0.2 in the multivariable analysis. For which we then performed backwards elimination, keeping only those with p<0.05, and comparing each subsequent model to the previous using a likelihood ratio test. Finally, we generated three separate models for each analysis (index and infected household members), including each contact parameter to the final model to assess the association with transmission.

### Ethics

The study protocol was approved by University of the Witwatersrand Human Research Ethics Committee (Reference M2008114). Participants in follow-up received grocery store vouchers of USD 3 per visit to compensate for time required for specimen collection and interview, and an additional voucher once proximity sensors were returned with no visible damage.

## Results

We screened 1,531 individuals and identified 277 (18%) positive for SARS-CoV-2, of which 124 (45%) were enrolled and included in the household cumulative infection risk analysis ^19^, with 373 household contacts. After data cleaning, we had contact data for 88 (71%) index cases and 252 (68%) household contacts (Supplementary Figure 1). Ninety-three individuals (19%, 36 index cases and 73 household contacts) were excluded due to non-compliance, where no contacts were logged, or sensors was stationary for the period based on accelerometer data. We were more likely to have contact data for individuals from the Soweto site, from larger households, and with no household member reporting smoking indoors (Supplementary Table 1). The median number of household members included in the analysis was 4 (inter quartile range [IQR] 3-5), with a median of 3 (IQR 1-4) SARS-CoV-2 cases per household and a median of 67% (IQR 50-100%) of household members infected (including index cases). Sixty-six percent (225/340) of individuals included in the analysis lived in a household with 3-5 members, and 49% (168/340) lived in a home with only 1-2 rooms used for sleeping, a third (53/340) living in households where crowding was reported (>2 people per sleeping room, Supplementary Table 1).

The overall median duration of daily close-range proximity events was 1,095 seconds (18 minutes, IQR 398-2,705 seconds), with a 39 second (IQR 32-49 seconds) average duration per contact event, and a median of 26 (IQR 10-58) close-range proximity events per day amongst household members (Figure 1, Supplementary Table 2). The highest median daily contact duration was observed between individuals within the <5 year, 5-12 year and 35-59 year groups (Figure 2 A, D). Similar patterns were also seen for median daily close-range proximity duration and frequency in children aged 5-12 and 13-17 years (Figure 2 B-F.

**Figure 1.**
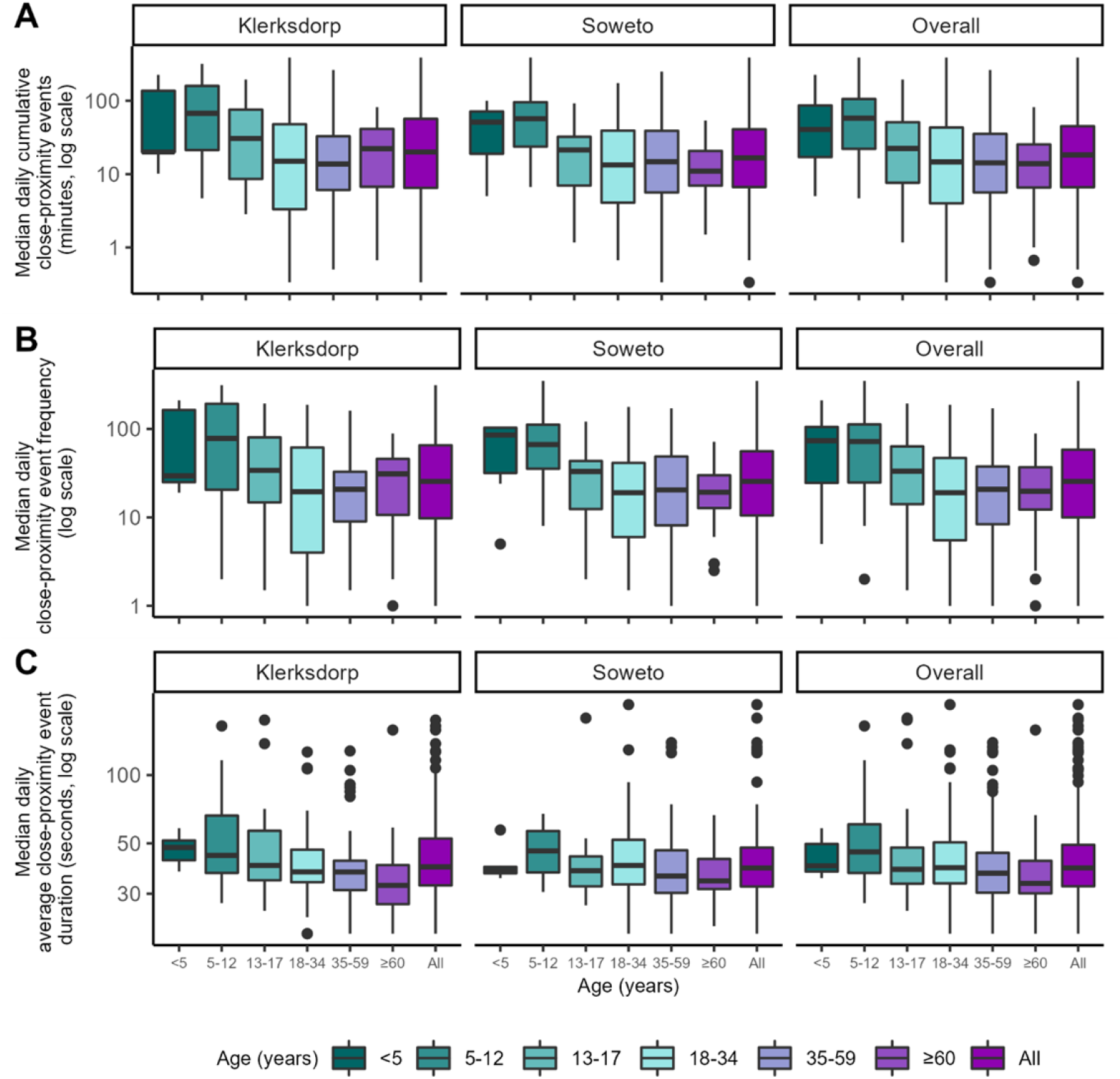
Contact parameters by age group (year) and site, Klerksdorp and Soweto, South Africa, September 2020 – October 2021. Horizontal line represents the median, box represents the 25^th^ and 75^th^ percentile, whiskers represent 1^st^ and 99^th^ percentile, circles indicate outliers.

**Figure 2.**
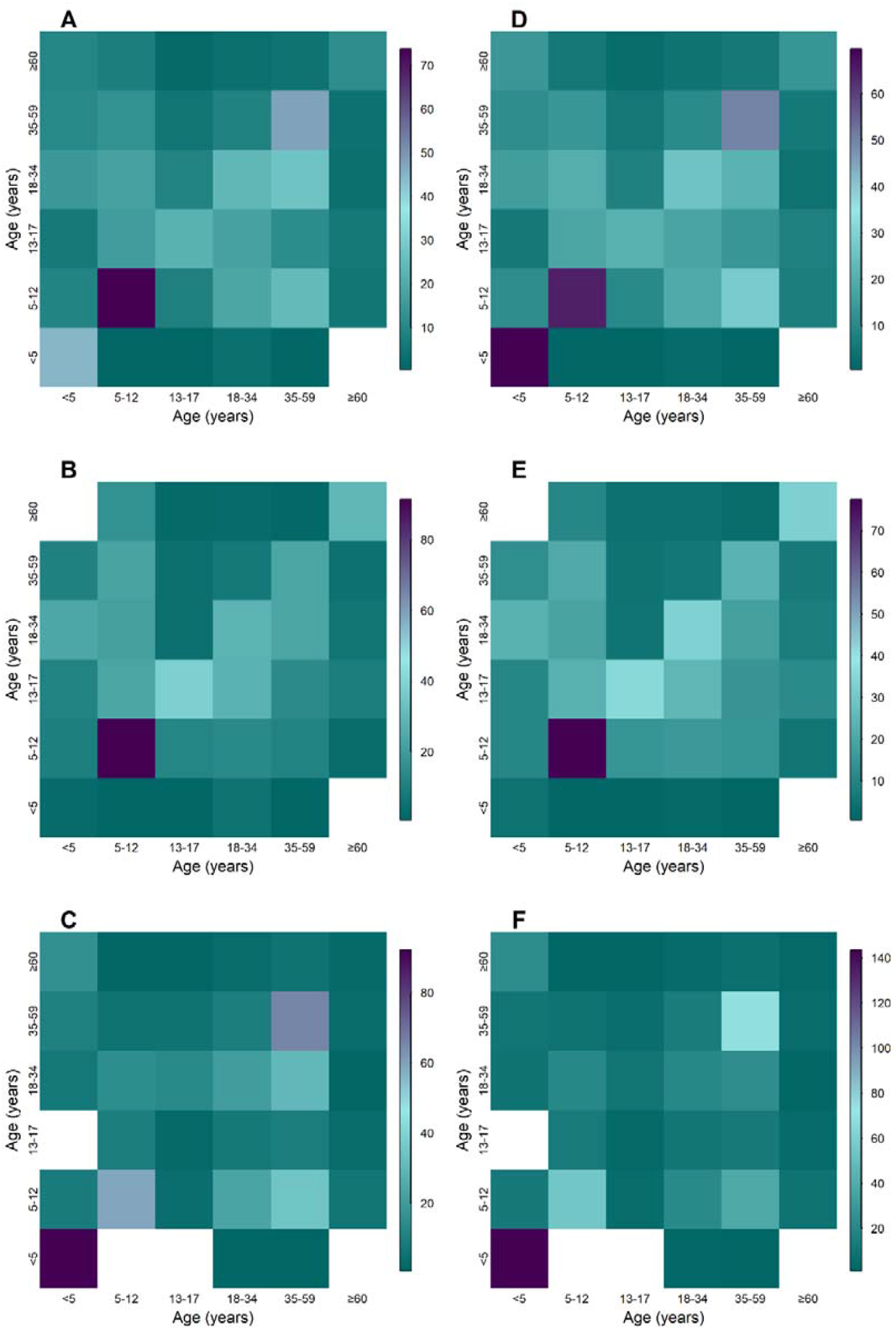
Aged-based contact matrices based close-proximity event duration (A-C) and frequency (D-E) for entire deployment period overall (A, D) Klerksdorp (B,D), and Soweto (C,F), September 2020 – October 2021. Teal denotes lowest value, purple highest, white no data for age group combination

We did not find any association between any of the contact parameters (either with the index case or all SARS-CoV-2 infected household members) and SARS-CoV-2 infection in the household using the Wilcoxon rank-sum test (p-values ranging 0.2-0.9, Table 1).

**Table 1.** Association of contact parameters with SARS-CoV-2 household acquisition* using the Wilcoxon rank-sum test, Klerksdorp and Soweto, South Africa, September 2020 – October 2021

| Contact parameter | p-value |
| --- | --- |
| Median daily contact duration with index | 0.82 |
| Median daily contact frequency with index | 0.71 |
| Median daily average contact duration with index | 0.22 |
| Median daily contact duration with infected household members | 0.25 |
| Median daily contact frequency with infected household members | 0.32 |
| Median daily average contact duration with infected household members | 0.95 |

\*Outcome investigated: testing positive for SARS-CoV-2

When assessing factors associated with SARS-CoV-2 transmission from presumptive index cases and acquisition in household members, none of the contact parameters were associated with SARS-CoV-2 transmission on univariate analysis. Sleeping in the same room as the index case was also not associated with transmission (OR 0.94, 95%CI 0.47 to 1.88). On multivariable analysis after controlling for index age and SARS-CoV-2 infecting variant, factors significantly associated with higher SARS-CoV-2 transmission and acquisition was index case minimum C_t_ value <30 (aOR 10.2 95%CI 1.4-77.4) compared to C_t_ >35, contacts aged 13-17 years (aOR 7.7 95%CI 1.0-58.2) compared to <5 years and female contacts (aOR 2.3 95%CI 1.1-4.8). Neither close-range proximity duration (aOR 1.0 95%CI 1.0-1.0), frequency (aOR 1.0 95%CI 1.0-1.0) or average close-range proximity duration (aOR 1.0 95%CI 1.0-1.0) with the index case were associated with acquisition (Table 2).

**Table 2.**
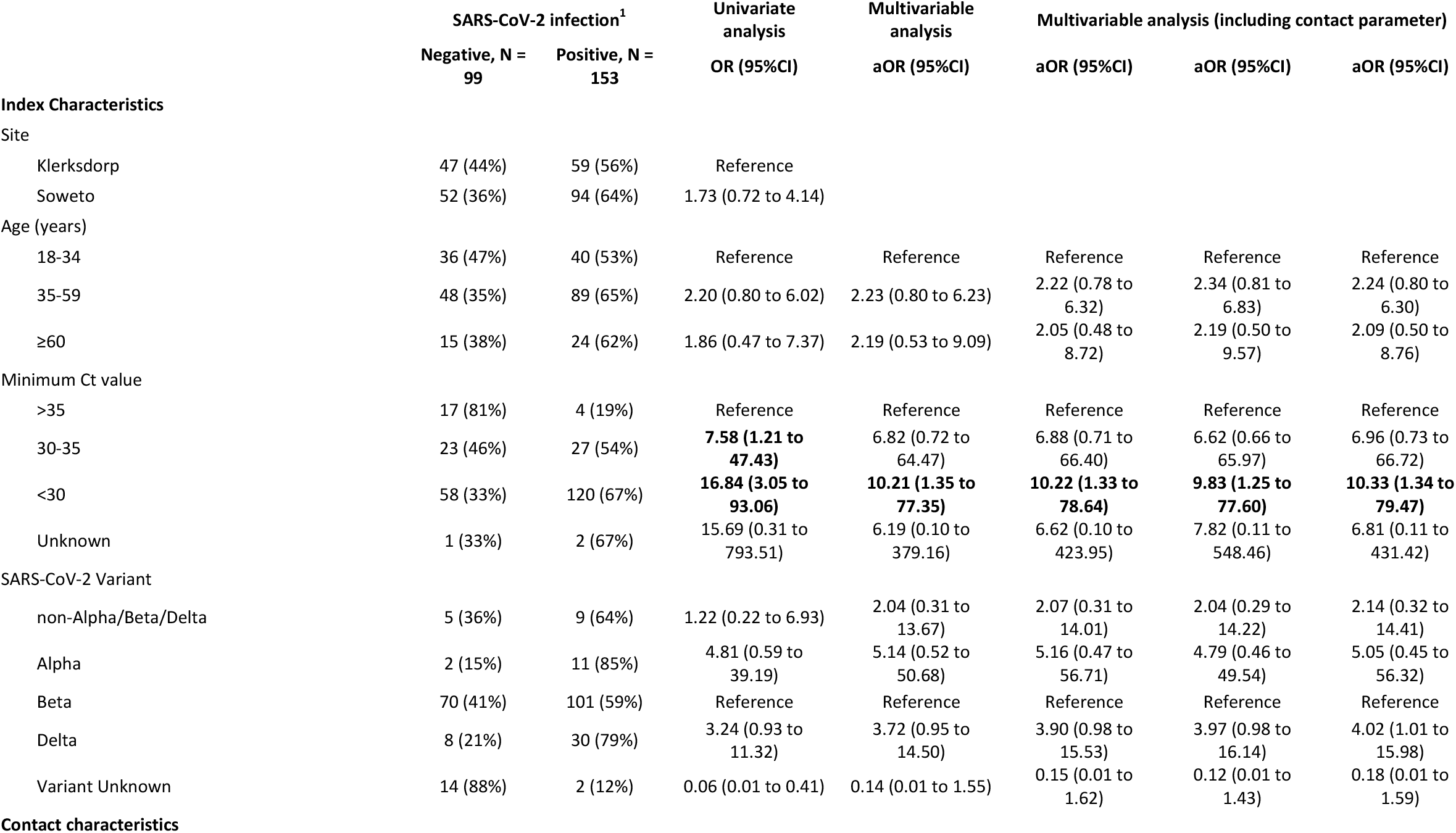
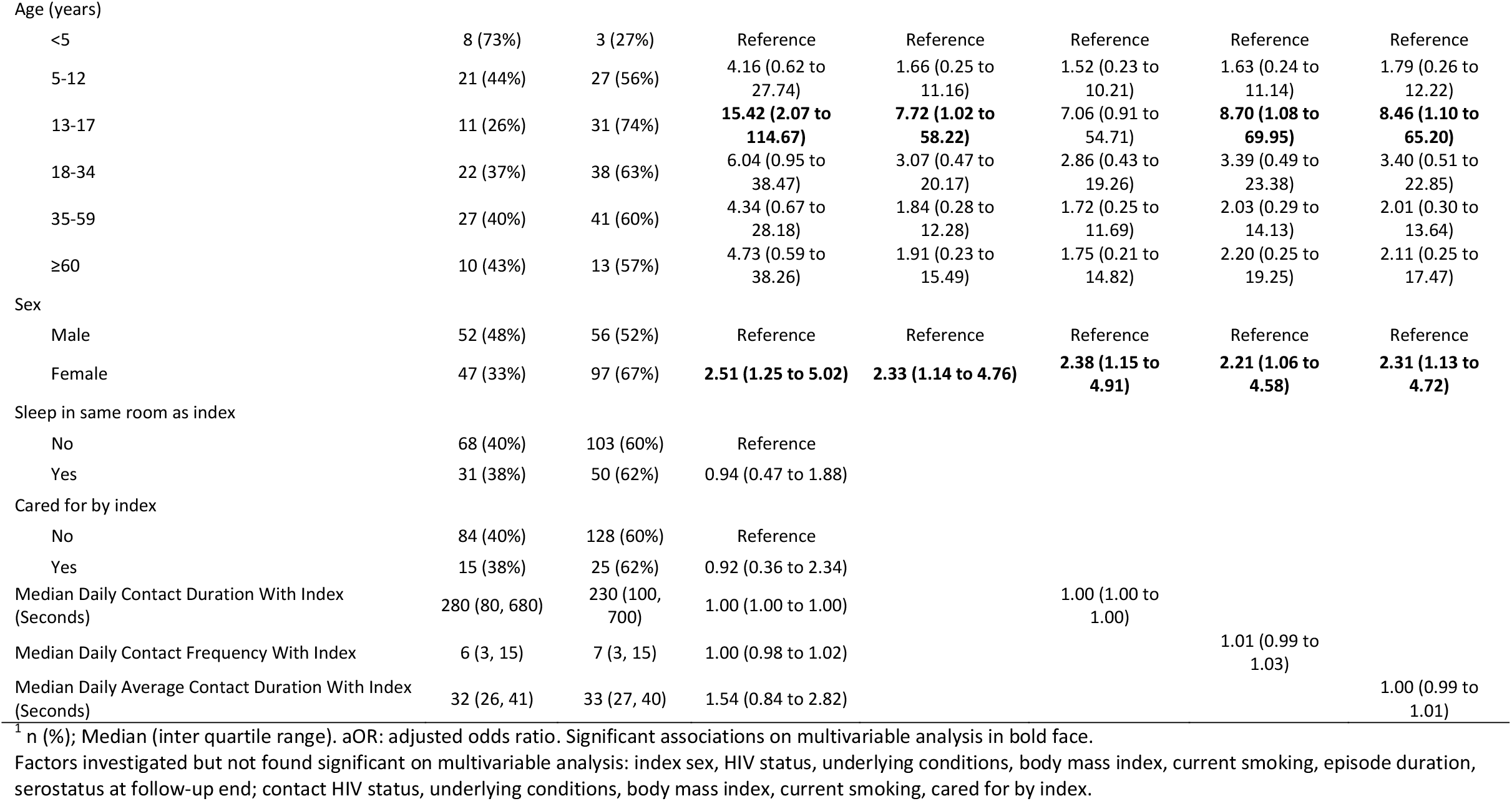
Factors associated with SARS-CoV-2 household transmission from index cases and acquisition in household contacts (contact parameters with index case), Klerksdorp and Soweto, South Africa, 2020-2021, (n=252)

When not considering transmission from the presumptive index case, but rather all SARS-CoV-2 infected household members, factors significantly associated with SARS-CoV-2 acquisition on multivariable analysis after controlling for SARS-CoV-2 infecting variant were contacts aged 13-17 years (aOR 9.4 95%CI 1.2-75.9), 18-34 years (aOR 10.1 95%CI 1.3-76.0) and 35-59 years (aOR 8.8 95%CI 1.1-67.7) compared to <5 years, being obese (aOR 4.1 95%CI 1.5-11.1) compared to normal weight, and not currently smoking (aOR 3.2 95%CI 1.2-9.2). Neither close-range proximity duration (aOR 1.0 95%CI 1.0-1.0), frequency (aOR 1.0 95%CI 1.0-1.0) or average close-range proximity duration (aOR 1.0 95%CI 1.0-1.0) with SARS-CoV-2 infected household members was associated with acquisition (Table 3).

**Table 3.**
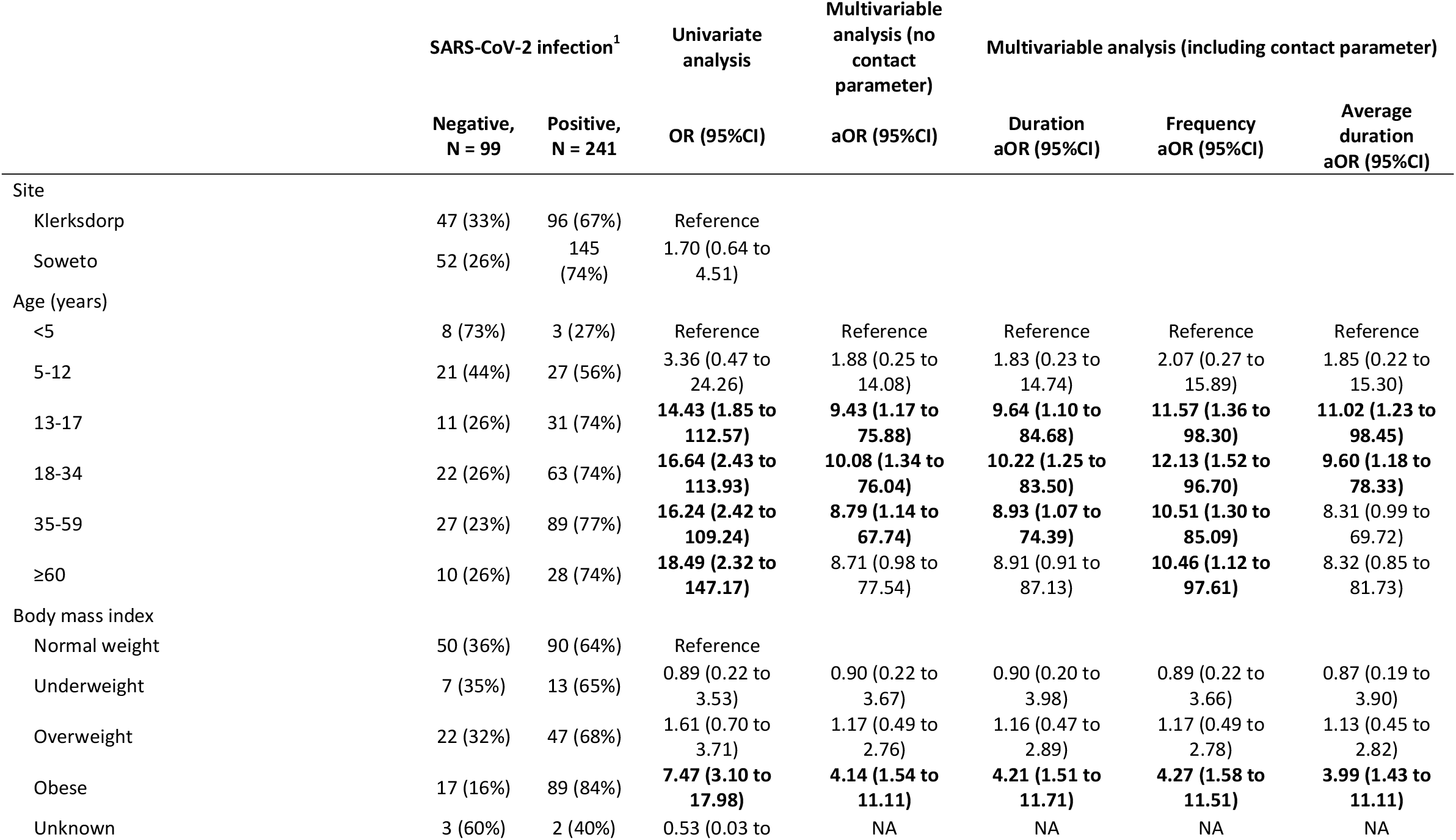
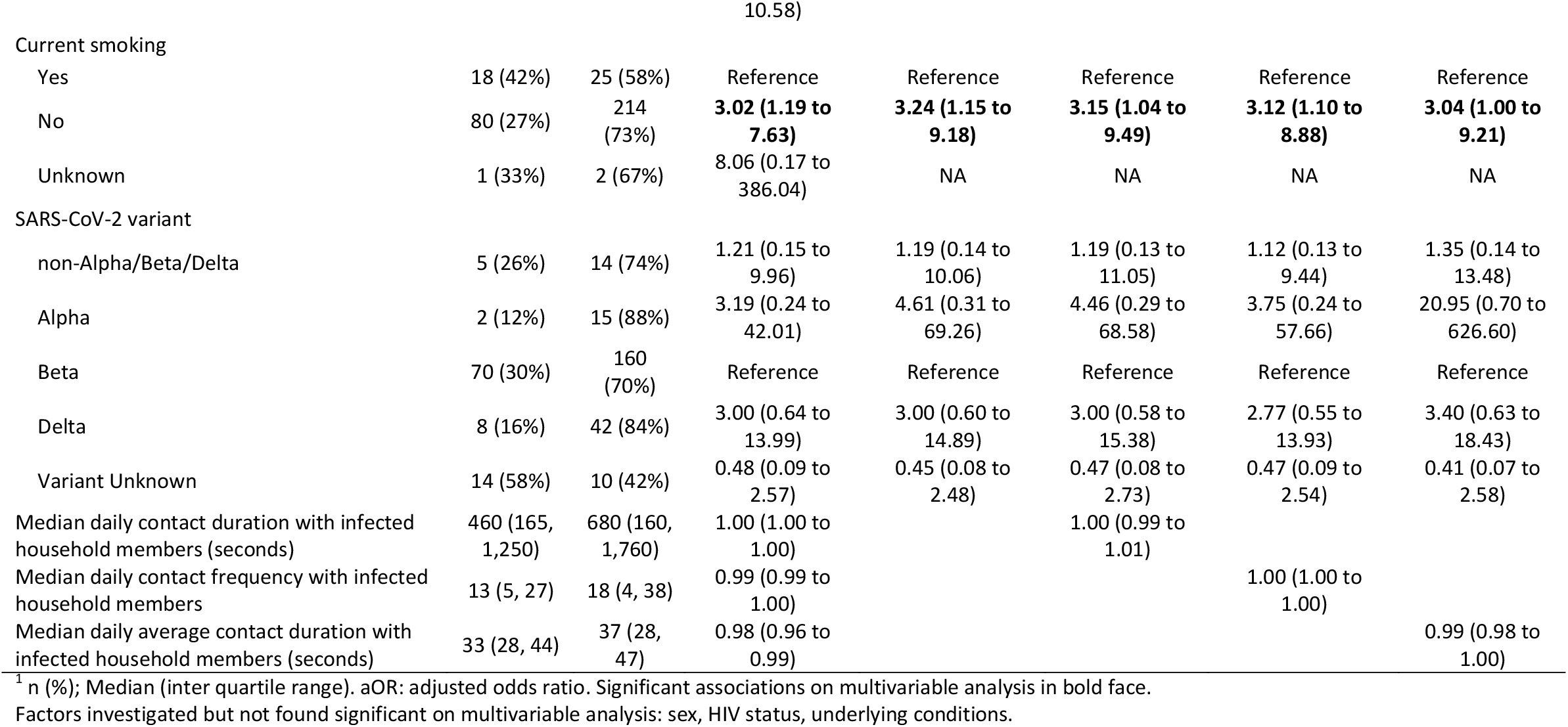
Factors associated with SARS-CoV-2 acquisition within in the household (contact parameters with SARS-CoV-2 infected household members), Klerksdorp and Soweto, South Africa, 2020-2021, (n=340)

## Discussion

In this case-ascertained, prospective household transmission study we did not find an association between the duration and frequency of close-range proximity events with SARS-CoV-2 infected household members with transmission in the household.

High-resolution contact patterns have been previously used to investigate influenza virus transmission routes in a hospital setting ^22^, and contact surveys to show the association between contacts, locations and influenza infection ^23^, the high correlation between pneumococcal infection risk and contact behaviour ^24^, and in the context of tuberculosis transmission where contact with adults are more important than contact with children ^25^. To our knowledge, there are few data available on the direct association of close-range proximity events and SARS-CoV-2, and none making use of high-resolution contact data. During contact tracing efforts early in the pandemic in Singapore, it was found that sharing a bedroom with an index case and speaking to the index case for more 30 minutes or longer increased the risk for infection ^26^. We did not see similar results when assessing sharing a bedroom with the index case, and this may be due to the already high level of crowding in included households. Although we observed an increase in infection risk with higher average contact durations with the index case on univariate analysis, this association was no longer seen when adjusting for age and other index and contact factors associated with transmission/acquisition.

There are several possible reasons why we did not observe an association with close-range proximity events and SARS-CoV-2 transmission, these can be classified as related to transmission dynamics or study limitations. One possibility is that along with droplet-mediated transmission during close-proximity contacts, airborne ^4,5^, and to a lesser extent, fomite-mediated transmission ^4^ may also play a role in the transmission of SARS-CoV-2 in the household. More evidence is becoming available showing that aerosol transmission may be a more important transmission route for SARS-CoV-2 than initially anticipated, especially so in poorly ventilated indoor environments^5,27^. Households in these communities do not have central air-conditioning or heating ^28^, and during the winter months ventilation may by poorer than in summer, although we did not measure this. Furthermore, sensors only measure face-to-face interactions, and if individuals were close to each other but not directly facing one another for extended durations, we would not have measured this, although sharing of the same air may have occurred. The ventilation within households should be considered in future studies, as this can be a target for intervention strategies to reduce secondary transmission. The high level of interaction in relatively crowded South African households may already be above the threshold for transmission risk, with host-characteristics like index viral load and contact age being more important to determine infection risk in this context. It is of interest that close-range proximity patterns within the household did not fully account for the differences in transmission based on age; with teenagers and adults experiencing the highest infection risk, but children aged 5-17 years having the highest contacts.

Our study had limitations both in design and execution. Due to the nature of the case-ascertained study design, we would have missed the period when the index case was most infectious, just before symptom onset ^4^, and the close-range contact patterns measured during the study may have been different after the household members were aware of the index SARS-CoV-2 case (leading to reduced contact), and again once secondary cases were informed of their infection status (leading to increased contact). We also did not collect any information on possible NPI usage in the households, like wearing of masks. A study from South Africa showed that individuals staying at home were less likely to wear a mask ^29^, but these data were not ascertained during a time when a household member was infected with SARS-CoV-2. We also did not consider where contacts took place (indoors or outdoors), which relates to ventilation and may have influenced transmission. We may have also misclassified the true index case if they were asymptomatic, and did not consider tertiary transmission chains in the index-directed analysis. To adjust for possible misclassification, we performed a grouped assessment investigating close-range proximity events with all SARS-CoV-2 infected household members. This grouped analysis may also have diluted possible associations with the true infector. We also did not consider multiple introductions within the household, although we did exclude households with more than one SARS-CoV-2 variant detected. During the peaks of waves of infection in South Africa, one variant was responsible for the majority of the infections ^30^, and the additional introductions within the household were likely to have been the same as the initial variant. Higher resolution sequencing data may be useful to more accurately identify chains of transmission within the household. Combining contact data with clinical and virological/bacteriological data have been shown to be useful to reconstruct transmission networks ^31^, and we will consider this for future analyses. Our measurement of close-range contact patterns was also limited by compliance, as during the cleaning process we identified 73 sensors that were not worn, based on accelerometer data. We also had limited data in some households, where some individuals did not consent to the contact aspect for the study, or where we were unable to retrieve data due to hardware failure, lost or damaged tags.

In conclusion, we did not observe an association with close-proximity contacts and SARS-CoV-2 transmission in the household. A case-ascertained, prospective household transmission study may not be well suited to investigate this question. A possible other study design to consider is randomly selected prospective household cohorts, but deployment of sensors for extended periods of time may be logistically challenging and lead to participant fatigue and households in a cohort may not experience infection episodes unless the community attack rate is very high. High-resolution contacts in other settings like schools or workplaces where contacts are less frequent could be useful to identify the type of contact events that may lead to SARS-CoV-2 transmission. It may be that aerosol transmission plays a more important role than droplet-mediated transmission, which would make ventilation within households can also be an important consideration for future studies, and increased ventilation can be a method to reduce secondary transmission in households. Nevertheless, our study provides high-resolution household contact data that can be used to parametrise future transmission models.

## Data Availability

The investigators welcome enquiries about possible collaborations and requests for access to the dataset. Data will be shared after approval of a proposal and with a signed data access agreement. Investigators interested in more details about this study, or in accessing these resources, should contact the corresponding author.

## Author contributions

Conception and design of study: JK, LD, LG, MT, SW, NM, AvG, NW, MM, CCo, CCa, ST

Data collection and laboratory processing: LM, JK, SW, LM, NM, AvG, NW

Analysis and interpretation: JK, LD, LG, MT, SW, CCo, CCa, ST

Accessed and verified underlying data: JK, LD, LG, MT, CCa

Drafted the Article: JK

All authors critically reviewed the Article. All authors had access to all the data reported in the study.

## Data availability

The contact network, selected individual characteristics and analysis script are available at https://github.com/crdm-nicd/sashts.git.

## Other members of the South African SARS-CoV-2 Household Transmission Study (SA-S-HTS) Group

Amelia Buys, ND (Centre for Respiratory Diseases and Meningitis, National Institute for Communicable Diseases of the National Health Laboratory Service, Johannesburg, South Africa.)

Daniel G. Amoako, PhD (Centre for Respiratory Diseases and Meningitis, National Institute for Communicable Diseases of the National Health Laboratory Service, Johannesburg, South Africa. School of Health Sciences, College of Health Sciences, University of KwaZulu-Natal, KwaZulu-Natal, South Africa.)

Dylan Toi, MD (Centre for Respiratory Diseases and Meningitis, National Institute for Communicable Diseases of the National Health Laboratory Service, Johannesburg, South Africa.)

Jinal N. Bhiman, PhD (Centre for Respiratory Diseases and Meningitis, National Institute for Communicable Diseases of the National Health Laboratory Service, Johannesburg, South Africa. School of Pathology, Faculty of Health Sciences, University of the Witwatersrand, Johannesburg, South Africa.)

Juanita Chewparsad, MSc (Perinatal HIV Research Unit (PHRU), University of the Witwatersrand, Johannesburg, South Africa.)

Kedibone Ndlangisa, PhD (Centre for Respiratory Diseases and Meningitis, National Institute for Communicable Diseases of the National Health Laboratory Service, Johannesburg, South Africa.)

Leisha Genade, MPH (Perinatal HIV Research Unit (PHRU), University of the Witwatersrand, Johannesburg, South Africa.)

Limakatso Lebina, PhD (Perinatal HIV Research Unit (PHRU), University of the Witwatersrand, Johannesburg, South Africa. Africa Health Research Institute (AHRI), Durban, South Africa)

Linda de Gouveia, ND (Centre for Respiratory Diseases and Meningitis, National Institute for Communicable Diseases of the National Health Laboratory Service, Johannesburg, South Africa.)

Mzimasi Neti, MPH (Centre for Respiratory Diseases and Meningitis, National Institute for Communicable Diseases of the National Health Laboratory Service, Johannesburg, South Africa.)

Retshidisitswe Kotane, BCMP (Centre for Respiratory Diseases and Meningitis, National Institute for Communicable Diseases of the National Health Laboratory Service, Johannesburg, South Africa.)

---

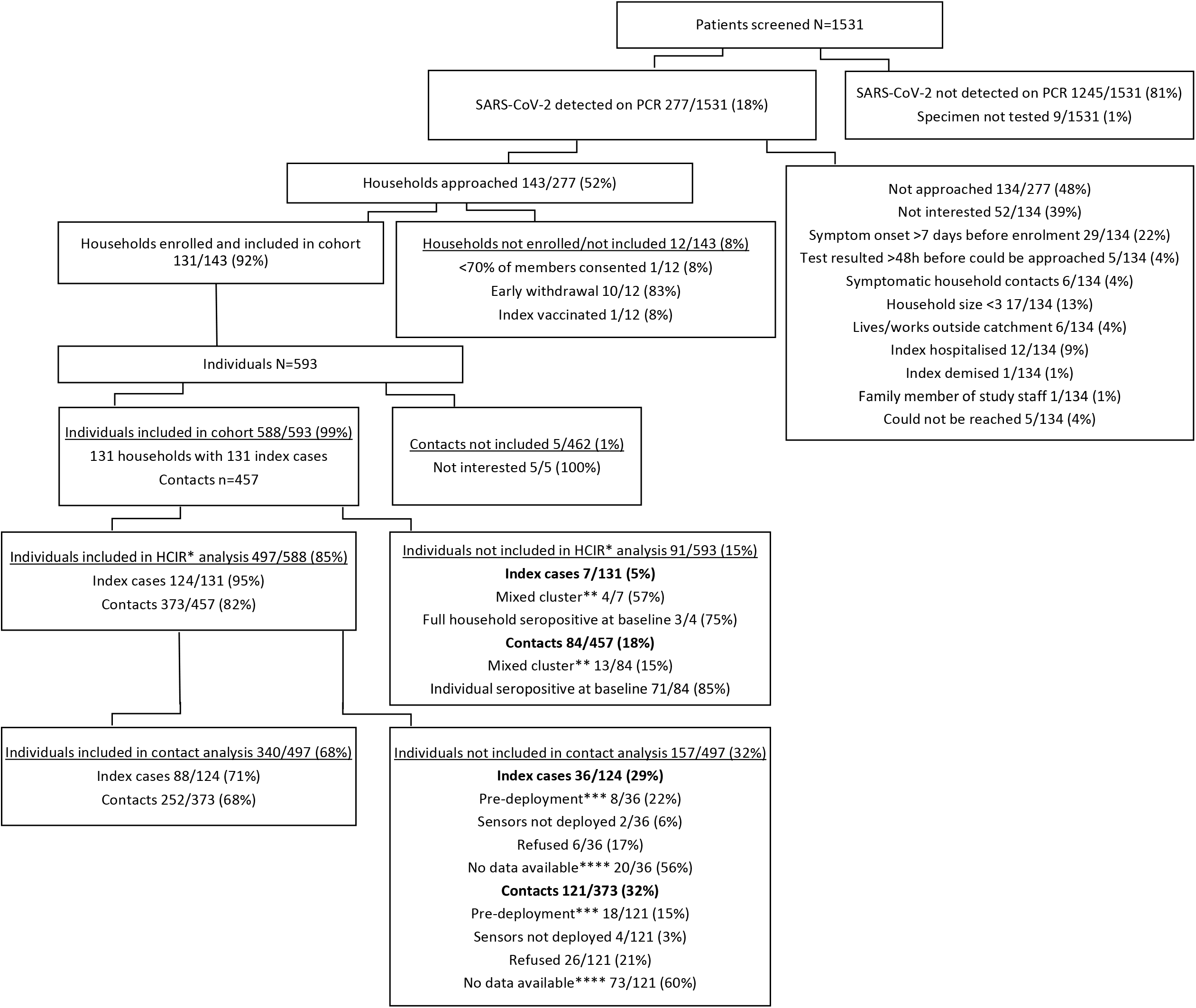

*Not included in HCIR analysis in study main manuscript (Kleynhans J, Walaza S, Martinson NA, et al. Household transmission of SARS-CoV-2 from adult index cases living with and without HIV in South Africa, 2020-2021: a case-ascertained, prospective observational household transmission study. Clin Infect Dis 2022.), **More than one SARS-CoV-2 variant detected in household, ***Household completed first week of follow-up before proximity sensors were deployed in field, ****No data available due to: Non-compliance based on lack of contacts logged or stationary accelerometer (n=73), Tag hardware failure (n=11), Tag lost/damaged (n=9)

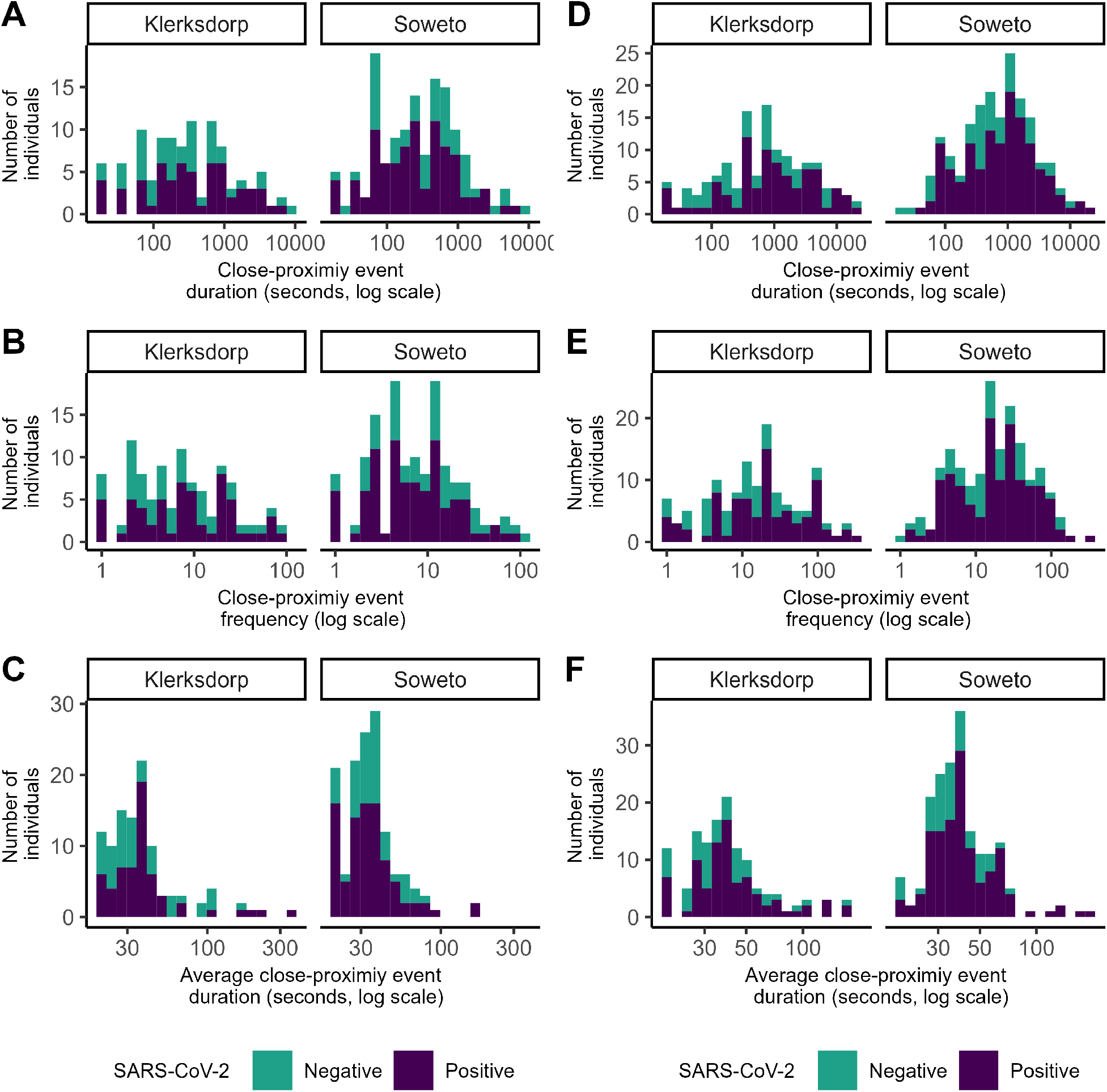

